# Risk Factor Stratification for Postoperative Delirium: A Retrospective Database Study

**DOI:** 10.1101/2022.03.06.22271982

**Authors:** Susana Vacas, Tristan Grogan, Drew Cheng, Ira Hofer

**Affiliations:** Department of Anesthesiology and Perioperative Medicine, David Geffen School of Medicine, University of California Los Angeles, Los Angeles, California, USA; Department of Medicine Statistics Core, David Geffen School of Medicine, University of California Los Angeles, Los Angeles, California, USA; Department of Anesthesiology and Medicine, Icahn School of Medicine at Mount Sinai

**Author notes:** <u>Corresponding author:</u> Susana Vacas, MD, PhD, 757 Westwood Plaza, Suite 2331, Los Angeles, CA, 90095.

**Keywords:** Postoperative delirium, risk factors, postoperative neurologic outcomes, perioperative neurocognitive disorders, surgery, anesthesia

## Abstract

**Background:** Postoperative Delirium (POD) is a disturbing reality for patients and their families. Absence of easy-to-use and accurate risk scores prompted us to retrospectively extract data from the electronic health records (EHR) to identify clinical factors associated with POD. We seek to create a multivariate nomogram to predict the risk of POD based upon the most significant clinical factors.

**Methods:** The EHR of patients >18 years of age undergoing surgery and had POD assessment were reviewed. Patient characteristics and study variables were summarized between delirium groups. We constructed univariate logistic regression models for POD using each study variable to estimate odds ratios (OR) and constructed a multivariable logistic regression model with stepwise variable selection. In order to create a clinically useful/implementable tool we created a nomogram to predict risk of delirium.

**Results:** Overall, we found a rate of POD of 3.7% across our study population. The Model achieved an AUC of the ROC curve of 0.83 (95% CI 0.82-0.84). We found that age, increased ASA score (ASA 3-4 OR 2.81, CI1.49-5.28, p<0.001), depression (OR 1.28, CI1.12-1.47, p<0.001), postoperative benzodiazepine use (OR 3.52, CI3.06-4.06, p<0.001) and urgent cases (Urgent OR 3.51, CI2.92-4.21, p<0.001; Emergent OR 3.99, CI3.21-4.96, p<0.001; Critically Emergent OR 5.30, CI3.53-7.96, p<0.001) were associated with POD.

**Conclusions:** We were able to distinguish the contribution of individual risk factors to the development of POD. We created a clinically useful easy-to-use tool that has the potential to accurately identify those at high-risk of delirium, a first step to prevent POD.

## Introduction

Each year approximately 52 million Americans undergo surgical procedures^1^, characterized by immense physiologic stress. Despite a fundamental goal of returning patients to optimal health, harmful cognitive changes, such as postoperative delirium (POD), are a common and well-established occurrence after surgery.^2^ POD is the most common surgical complication in older adults^3^ and is estimated to cost $32.9 billion per year in the US alone.^4–6^

POD doubles a patient’s risk of post-discharge institutionalization, enhances all-cause mortality by 10-20% for every 48 hours of delirium,^7^ and is linked to long-term cognitive decline, including a 8-fold increased risk of dementia.^8–10^ It is estimated that 30-40% of POD cases (and ultimately long-lasting deficits) are preventable.^2,11,12^

Currently, perioperative prevention strategies are resource and personnel intensive and general prevention for all surgical patients is not cost-effective and probably unnecessary. A more realistic solution is risk stratification, whereby patients who are at high risk for POD can be targeted for preventative measures. Recognizing which patients are at heightened risk for developing POD continues to be an active area of research. Although several risk factors have been identified to be predictors of POD, most studies are limited by small sample size and/or are targeted for specific procedures or patient groups.^13,14–18^ By 2014, a systemic review and metanalysis identified 37 different risk prediction models for POD, but most were not externally validated.^15,19^ Approximately 80% of these tools focused on cardiac and orthopaedic surgery,^19,20^ while some were limited to a single type of pathology.^13^ Unfortunately, many commonly-used POD risk stratification instruments show no superior advantage in application and performance than random chance,^14,15,21^ which may be indicative of the small-sized studies on which these instruments were developed.

Over the past several years, the introduction of electronic health records (EHR) has increased the feasibility of obtaining a number of data points across a large patient population. In particular, our group has demonstrated the ability to extract complex medical phenotypes from a variety of EHR data.^22,23^ The ability to extract these data from the EHR allows for the possibility of expanding on current work on POD risk stratification to include larger populations and a wider variety of surgical procedures.

In this manuscript we seek to leverage our expertise in complex data extraction from the EHR to retrospectively identify clinical factors associated with the risk of POD. As a secondary outcome we then seek to create a multivariate nomogram to predict the risk of POD based upon the most significant clinical factors.

## Methods

### Data Extraction

The University of California Los Angeles (UCLA) Institutional Review Board approved and waived the requirement for informed consent (IRB#15-000518) for this single-center retrospective observational study. All patients aged 18 years and older undergoing surgery or procedures under anesthesia between April 2013 and July 2020 at the UCLA Medical Center and affiliated surgery centers were considered eligible for inclusion. We excluded from the analysis patients aged less than 18 years old, and cases that spent less than one night in the hospital.

All data for this study were extracted from the Perioperative Data Warehouse (PDW), a custom-built robust data warehouse containing all patients who have undergone surgery at the UCLA Health since the implementation of our EHR (EPIC Systems, Madison, WI) in March 2013. We have previously described the creation of the PDW, which has a 3-stage design.^24,25^ Briefly, in the first stage, data are extracted from EPIC’s Clarity database into 29 tables organized around 3 distinct concepts: patients, surgical procedures, and health system encounters. These data are then cleaned and used to populate a series of 4000 distinct measures and metrics such as procedure duration, readmissions, admission International Classification of Diseases (ICD) codes, and postoperative outcomes.

### Definition of Outcome Measure: Postoperative Delirium

We defined the primary outcome as POD using the Confusion Assessment Method (CAM) score.^24^ The CAM is a commonly used tool that identifies the presence/absence of POD and is consistently applied and documented in the EHR of surgical patients at UCLA.^24^ It consists of a series of yes/no questions meeting the Diagnostic and Statistical Manual of Mental Disorders (DSM-5) criteria for delirium: acute onset and/or fluctuating course during the day of disturbance of attention, awareness or cognition not explained by another preexisting or established neurocognitive disorder.^25,26^ The responses to the individual metrics are stored in the EHR at the time of documentation. These data were extracted from the EHR and the individual metrics grouped at the level of the calendar day. For that calendar day, a positive response to any of the CAM questions was taken as the patient having POD on that day. For our study, a patient was considered to have POD if they had a single positive CAM score in the seven calendar days following surgery. Because not all elements were always documented contemporaneously, in the even that only some elements were charted the most recent values for the other elements were used to compute the score.^25,26^ Patients who were never screened for delirium were excluded from the study.

### Perioperative characteristics

For the purposes of this study, preoperative patient characteristics were chosen based on current literature and included age, sex, height, weight, American Society of Anesthesiologists (ASA) status, surgical service, procedure type, inpatient versus outpatient status, elective versus emergent/critically emergent procedure, metabolic equivalents of task, SPICES score and comorbidities such as congestive heart failure, peripheral vascular disease, pulmonary history including Chronic Obstructive pulmonary Disease, kidney and endocrine disease, obstructive sleep apnea, neuropsychiatric history including anxiety and depression, history of tobacco or alcohol use, as well as imaging (echocardiogram) and laboratory values (glomerular filtration rate).

Intraoperative characteristics included type of anesthetic, duration of anesthesia, administration of benzodiazepines and intraoperative hemodynamic monitoring data, such as time spent with mean arterial pressure < 55 or <65 mmHg.

### Statistical Analysis

Patient characteristics and study variables were summarized between delirium groups using frequency (%) or mean (SD) and compared between groups using the chi-square test or t-test as appropriate. We then constructed univariate logistic regression models for delirium using each study variable to estimate odds ratios (OR) with 95% confidence intervals. Next, we constructed a multivariable logistic regression model with stepwise variable selection to get a parsimonious model. Finally, in order to create a more clinically useful/implementable tool we created a nomogram from this model to predict risk of delirium both visually and as a point system score. The overall discrimination/prognostic ability of the model was assessed using the area under the ROC curve (AUC). P-values <0.05 were considered statistically significant and statistical analyses were performed using IBM SPSS V26 (Armonk, NY) and R V4.1.0 (www.r-project.org, Vienna, AU).

## Results

### Patient Demographics and Univariate Analysis

Table 1 shows the overall demographics of the cohort and the rates of the covariates studied. We identified 32,734 patients who met inclusion criteria. The overall incidence of POD was 3.7%. The cohort consisted of 7,315 (22.3%) patients aged 65-75 years and 5,401 (16.5%) patients age >75 years old. The majority (20,199 (62.7%)) of patients were ASA physical status class 3 or 4 and the most common surgical services were Orthopedics (7,200), General Surgery (6,126), and Neurosurgery (3,345). Most cases (19,950 (60.9%)) were elective.

**Table 1.** Overall cohort demographics and univariate analysis of risk factors and delirium. P-values listed are those of the logistic regression for individual risk factors. Values presented as frequency (%) unless otherwise noted; *values presented as mean / SD

| Demographic / clinical characteristics | No Delirium<br>31539 96,3% |  | Delirium<br>1195 3,7% |  | Total<br>32 734 100,0% |  | Delirium Incidence | Univariate OR (95% CI) | p-value Log. Reg. |
| --- | --- | --- | --- | --- | --- | --- | --- | --- | --- |
| **AGE GROUP** |  |  |  |  |  |  |  |  | <0.001 |
| 18-45 | 7522 | 23,8% | 98 | 8,2% | 7 620 | 23,3% | 1,3% | REF |  |
| 45-65 | 12055 | 38,2% | 343 | 28,7% | 12 398 | 37,9% | 2,8% | 2.18 (1.74-2.74) | <0.001 |
| 65-75 | 7075 | 22,4% | 240 | 20,1% | 7 315 | 22,3% | 3,3% | 2.60 (2.05-3.30) | <0.001 |
| >75 | 4887 | 15,5% | 514 | 43,0% | 5 401 | 16,5% | 9,5% | 8.07 (6.49-10.05) | <0.001 |
| **ASA_SCORE Group** |  |  |  |  |  |  |  |  | <0.001 |
| 1-2 | 11783 | 37,4% | 102 | 8,5% | 11 885 | 36,3% | 0,9% | REF |  |
| 3-4 | 19139 | 60,7% | 1060 | 88,7% | 20 199 | 61,7% | 5,2% | 6.40 (5.22-7.85) | <0.001 |
| 5 | 131 | 0,4% | 17 | 1,4% | 148 | 0,5% | 11,5% | 14.99 (8.72-25.77) | <0.001 |
| **PRIM_SURG_SERVICE** |  |  |  |  |  |  |  |  | <0.001 |
| Obstetrics and Gynecology | 1476 | 4,7% | 7 | 0,6% | 1 483 | 4,5% | 0,5% | REF |  |
| Cardiac Surgery | 1444 | 4,6% | 48 | 4,0% | 1 492 | 4,6% | 3,2% | 7.01 (3.16-15.54) | <0.001 |
| Gastroenterology | 1259 | 4,0% | 75 | 6,3% | 1 334 | 4,1% | 5,6% | 12.56 (5.77-27.35) | <0.001 |
| General Surgery | 5976 | 18,9% | 150 | 12,6% | 6 126 | 18,7% | 2,4% | 5.29 (2.48-11.32) | <0.001 |
| Liver Transplant | 771 | 2,4% | 68 | 5,7% | 839 | 2,6% | 8,1% | 18.60 (8.50-40.69) | <0.001 |
| Neurosurgery | 3142 | 10,0% | 203 | 17,0% | 3 345 | 10,2% | 6,1% | 13.62 (6.40-29.01) | <0.001 |
| Orthopaedics | 6825 | 21,6% | 375 | 31,4% | 7 200 | 22,0% | 5,2% | 11.59 (5.47-24.52) | <0.001 |
| Other | 1053 | 3,3% | 43 | 0,1% | 1 096 | 3,3% | 3,9% | 8.61 (3.86-19.22) | <0.001 |
| Otolaryngology | 2413 | 7,7% | 41 | 3,4% | 2 454 | 7,5% | 1,7% | 3.58 (1.60-8.01) | 0.002 |
| Plastic Surgery | 979 | 3,1% | 18 | 1,5% | 997 | 3,0% | 1,8% | 3.88 (1.61-9.32) | 0.002 |
| Surgical Oncology | 637 | 2,0% | 17 | 1,4% | 654 | 2,0% | 2,6% | 5.63 (2.32-13.64) | <0.001 |
| Thoracic Surgery | 823 | 2,6% | 26 | 2,2% | 849 | 2,6% | 3,1% | 6.66 (2.88-15.41) | <0.001 |
| Urology | 3593 | 11,4% | 65 | 5,4% | 3 658 | 11,2% | 1,8% | 3.82 (1.75-8.34) | 0,001 |
| Vascular Surgery | 743 | 2,4% | 30 | 2,5% | 773 | 2,4% | 3,9% | 8.51 (3.72-19.47) | <0.001 |
| **CASE_TYPE** |  |  |  |  |  |  |  |  | <0.001 |
| Elective | 19656 | 62,3% | 294 | 24,6% | 19 950 | 60,9% | 1,5% | REF |  |
| Critically Emergent | 343 | 1,1% | 39 | 3,3% | 382 | 1,2% | 10,2% | 7.60 (5.35-10.80) | <0.001 |
| Emergent | 2306 | 7,3% | 182 | 15,2% | 2 488 | 7,6% | 7,3% | 5.28 (4.36-6.38) | <0.001 |
| Inpatient | 2886 | 9,2% | 261 | 21,8% | 3 147 | 9,6% | 8,3% | 6.05 (5.10-7.18) | <0.001 |
| Transplant | 1226 | 3,9% | 37 | 3,1% | 1 263 | 3,9% | 2,9% | 2.02 (1.43-2.85) | <0.001 |
| Urgent | 4310 | 13,7% | 348 | 29,1% | 4 658 | 14,2% | 7,5% | 5.40 (4.61-6.33) | <0.001 |
| **Anesthesia duration\*** | 244,1 | 149,9 | 243,1 | 167,7 |  |  |  | 1.00 (1.00-1.00) | 0,835 |
| **ANESTHESIA_TYPE** |  |  |  |  |  |  |  |  | 0,584 |
| General | 27422 | 86,9% | 1049 | 87,8% | 28 471 | 87% | 3,7% | REF |  |
| Epidural | 366 | 1,2% | 11 | 0,9% | 377 | 1% | 2,9% | 1.25 (0.81-1.94) | 0,313 |
| MAC | 2650 | 8,4% | 91 | 7,6% | 2 741 | 8% | 3,3% | 0.99 (0.47-2.07) | 0,967 |
| Regional Block | 145 | 0,5% | 7 | 0,6% | 152 | 0% | 4,6% | 1.13 (0.70-1.82) | 0,632 |
| Spinal | 688 | 2,2% | 21 | 1,8% | 709 | 2% | 3,0% | 1.58 (0.66-3.79) | 0,304 |
| **Male sex (ref=female)** | 15310 | 48,5% | 654 | 54,7% | 15 964 | 49% | 4,1% | 0.78 (0.70-0.88) | <0.001 |
| **BMI** |  |  |  |  |  |  |  |  | <0.001 |
| 18.5-29.99 | 21228 | 67,3% | 841 | 70,4% | 22 069 | 67% | 3,8% | REF |  |
| < 18.5 | 1250 | 4,0% | 107 | 9,0% | 1 357 | 4% | 7,9% | 2.16 (1.75-2.66) | <0.001 |
| 30-39.99 | 7189 | 22,8% | 202 | 16,9% | 7 391 | 23% | 2,7% | 0.71 (0.61-0.83) | <0.001 |
| 40+ | 1571 | 5,0% | 26 | 2,2% | 1 597 | 5% | 1,6% | 0.42 (0.28-0.62) | <0.001 |
| **Positive SPICES Preoperatively** | 30785 | 97,6% | 1169 | 97,8% | 31 954 | 98% | 3,7% | 1.10 (0.74-1.64) | 0,633 |
| **History of Tobacco Use** | 1659 | 5,3% | 74 | 6,2% | 1 733 | 5% | 4,3% | 1.28 (1.00-1.63) | 0,047 |
| **History of Alcohol Use** | 13976 | 44,3% | 360 | 30,1% | 14 336 | 44% | 2,5% | 0.58 (0.51-0.66) | <0.001 |
| History of Depression | 7994 | 25,3% | 394 | 33,0% | 8 388 | 26% | 4,7% | 1.45 (1.28-1.64) | <0.001 |
| History of Anxiety | 5400 | 17,1% | 154 | 12,9% | 5 554 | 17% | 2,8% | 0.72 (0.60-0.85) | <0.001 |
| History of Obstructive Sleep Apnea | 2839 | 9,0% | 124 | 10,4% | 2 963 | 9% | 4,2% | 1.17 (0.97-1.42) | 0,104 |
| History of Diabetes | 8446 | 26,8% | 414 | 34,6% | 8 860 | 27% | 4,7% | 1.45 (1.28-1.64) | <0.001 |
| History of Congestive Heart Failure | 2379 | 7,5% | 197 | 16,5% | 2 576 | 8% | 7,6% | 2.42 (2.07-2.84) | <0.001 |
| History of Peripheral Vascular Disease | 3314 | 10,5% | 222 | 18,6% | 3 536 | 11% | 6,3% | 1.94 (1.67-2.26) | <0.001 |
| History of COPD | 5564 | 17,6% | 255 | 21,3% | 5 819 | 18% | 4,4% | 1.27 (1.10-1.46) | 0,001 |
| **PREOP_METS** |  |  |  |  |  |  |  |  | <0.001 |
| Excellent | 2297 | 7,3% | 19 | 1,6% | 2 316 | 7% | 0,8% | REF |  |
| Good | 11874 | 37,6% | 250 | 20,9% | 12 124 | 37% | 2,1% | 2.55 (1.59-4.07) | <0.001 |
| Poor | 2450 | 7,8% | 225 | 18,8% | 2 675 | 8% | 8,4% | 11.10 (6.93-17.79) | <0.001 |
| Unable to assess | 510 | 1,6% | 56 | 4,7% | 566 | 2% | 9,9% | 13.28 (7.82-22.53) | <0.001 |
| Preoperative ejection fraction (n=4,309 , 339)* | 62,6 | 12,60 | 63,6 | 13,90 |  |  |  | 1.01 (1.00-1.02) | 0,161 |
| Glomerular filtration Rate* | 88,5 | 42,50 | 86,4 | 51,20 |  |  |  | 1.00 (1.00-1.00) | 0,108 |
| Premedication with Midazolam | 16952 | 53,7% | 357 | 29,9% | 17 309 | 53% | 2,1% | 0.37 (0.32-0.42) | <0.001 |
| Perioperative Precedex Use | 623 | 2,0% | 22 | 1,8% | 645 | 2% | 3,4% | 0.93 (0.61-1.43) | 0,743 |
| Perioperative Ketamine Use | 0 | 0,0% | 0 | 0,0% | - | 0% | -- | -- | -- |
| Postoperative Benzodiazepine use | 4289 | 13,6% | 467 | 39,1% | 4 756 | 15% | 9,8% | 4.08 (3.61-4.60) | <0.001 |
| Duration MAP<65* | 31,5 | 76,90 | 35,7 | 55,0 |  |  |  | 1.00 (1.00-1.01) | 0,111 |
| Duration MAP<55* | 5,5 | 21,0 | 7,8 | 24,0 |  |  |  | 1.00 (1.00-1.01) | 0,001 |

Overall, in univariate analysis a wide variety of factors were directly associated with POD. Of note, patient age (45-65 years old, OR 2.18, 95% CI 1.74-2.74, p <0.001; 65-75 years old, OR 2.60, 95% CI 2.05-3.30, p <0.001; >75 years old, OR 8.07, 95% CI 6.49-10.05, p <0.001), ASA physical status (ASA 3-4 OR 6.40, 95% CI 5.22-7.85, p <0.001), female sex, surgical service (Cardiac surgery OR 7.01, 95% CI 3.16-15.54, p <0.001; Gastroenterology OR 12.56, 95% CI 5.77-27.35, p <0.001; General Surgery OR 5.29, 95% CI 2.48-11.32, p <0.001; Liver Transplant OR 18.60, 95% CI 8.50-40.69, p <0.001; Neurosurgery OR 13.62, 95% CI 6.40-29.01, p <0.001; Orthopaedics OR 11.59, 95% CI 5.47-24.52, p <0.001; Otolaryngology OR 3.58, 95% CI 1.60-8.01, p =0.002; Plastic Surgery OR 3.88, 95% CI 1.61-9.32, p =0.002; Surgical Oncology OR 5.63, 95% CI 2.32-13.64, p <0.001; Thoracic Surgery OR 6.66, 95% CI 2.88-15.41, p <0.001; Urology OR 3.82, 95% CI 1.75-8.34, p <0.001; Vascular Surgery OR 8.51, 95% CI3.72-19.47, p <0.001) and case urgency (Urgent OR 5.40, 95% CI 4.61-6.33, p <0.001; Emergent OR 5.28, 95% CI 4.36-6.38, p <0.001; Critically Emergent OR 7.60, 95% CI 5.35-10.80, p <0.001; Inpatient OR 6.05, 95% CI 5.10-7.18, p <0.001; Transplant OR 2.02, 95% CI 1.43-2.85, p <0.001) were associated with POD. In addition, BMI<18.5 (OR 2.16, 95% CI 1.75-2.66, p <0.001), BMI 30-39.99 (OR 0.71, 95% CI 0.61-0.83, p <0.001), BMI >40 (OR 0.42, 95% CI 0.28-0.62, p <0.001), depression (OR 1.45, 95% CI 1.28-1.64, p <0.001) and postoperative benzodiazepine use (OR 4.08, 95% CI 3.61-4.60, p <0.001) were associated with POD; however, premedication with benzodiazepines was not associated with POD, nor was anesthesia type. Further variables study can be found in Table 1.

### Multivariate Model

Table 2 shows the odds ratios for the features included in the multivariate analysis of POD. Overall, the Model achieved an AUC of the ROC curve of 0.83 (95% CI 0.82-0.84). Age above 45 was consistently associated with POD, with age greater than 75 years having a OR 8.26, 95% CI 6.39-10.68 of POD. In addition, increased ASA physical status score (ASA 3-4 OR 2.81, 95% CI 1.49-5.28, p <0.001), history of depression (OR 1.28, 95% CI 1.12-1.47, p <0.001), postoperative benzodiazepine use (OR 3.52, 95% CI 3.06-4.06, p <0.001) and more urgent cases (Urgent OR 3.51, 95% CI 2.92-4.21, p <0.001; Emergent OR 3.99, 95% CI 3.21-4.96, p <0.001; Critically Emergent OR 5.30, 95% CI 3.53-7.96, p <0.001) were associated with POD. Of interest premedication with benzodiazepines was protective of POD.

**Table 2.** Multivariate odds ratios for postoperative delirium and p-values for each category

| Demographic / clinical characteristics | Multivariable OR (95% CI) | p-value |
| --- | --- | --- |
| **Age group** |  | <0.001 |
| 18-45 (ref) | -- |  |
| 45-65 | 2.39 (1.87-3.06) |  |
| 65-75 | 3.26 (2.51-4.22) |  |
| >75 | 8.26 (6.39-10.68) |  |
| **ASA score group** |  | <0.001 |
| 1-2 (ref) | -- |  |
| 3-4 | 2.86 (2.29-3.58) |  |
| 5 | 2.81 (1.49-5.28) |  |
| **Male sex (ref=female)** | 1.29 (1.13-1.48) | <0.001 |
| **Hx Depression** | 1.28 (1.12-1.47) | <0.001 |
| **Perioperative midazolam** | 0.75 (0.65-0.87) | <0.001 |
| **Postoperative benzodiazepine** | 3.52 (3.06-4.06) | <0.001 |
| **Primary Surgical Service** |  | <0.001 |
| Obstetrics and Gynecology (ref) | -- |  |
| Cardiac Surgery | 1.15 (0.48-2.76) |  |
| Gastroenterology | 1.01 (0.43-2.39) |  |
| General Surgery | 1.41 (0.61-3.25) |  |
| Liver Transplant | 3.10 (1.30-7.43) |  |
| Neurosurgery | 4.50 (1.96-10.34) |  |
| Orthopaedics | 2.86 (1.25-6.52) |  |
| Other | 1.35 (0.55-3.28) |  |
| Otolaryngology | 1.20 (0.50-2.90) |  |
| Plastic Surgery | 2.13 (0.82-5.53) |  |
| Surgical Oncology | 1.17 (0.44-3.07) |  |
| Thoracic Surgery | 1.85 (0.74-4.62) |  |
| Urology | 1.37 (0.58-3.25) |  |
| Vascular Surgery | 1.66 (0.67-4.10) |  |
| **BMI categories** |  | 0,001 |
| 18.5-29.99 (ref) | -- |  |
| < 18.5 | 1.45 (1.15-1.84) |  |
| 30-39.99 | 0.93 (0.78-1.10) |  |
| 40+ | 0.66 (0.44-0.99) |  |
| **CASE_TYPE** |  | <0.001 |
| ELECTIVE (ref) | -- |  |
| CRITICALLY EMERGENT | 5.30 (3.53-7.96) |  |
| EMERGENT | 3.99 (3.21-4.96) |  |
| INPATIENT | 3.49 (2.89-4.23) |  |
| TRANSPLANT CASE | 1.88 (1.25-2.83) |  |
| URGENT | 3.51 (2.92-4.21) |  |
| **AUC (95% CI)** | 0.85 (0.84-0.86) |  |

### Creation of a score for POD risk stratification

In order to facilitate prospective risk stratification Figure 1 contains a nomogram that can be used to convert the odds ratios from the multivariate model into a “score” for POD prediction. The points associated with the various sections of the nomogram can be found in supplementary table 1.

**Figure 1.**
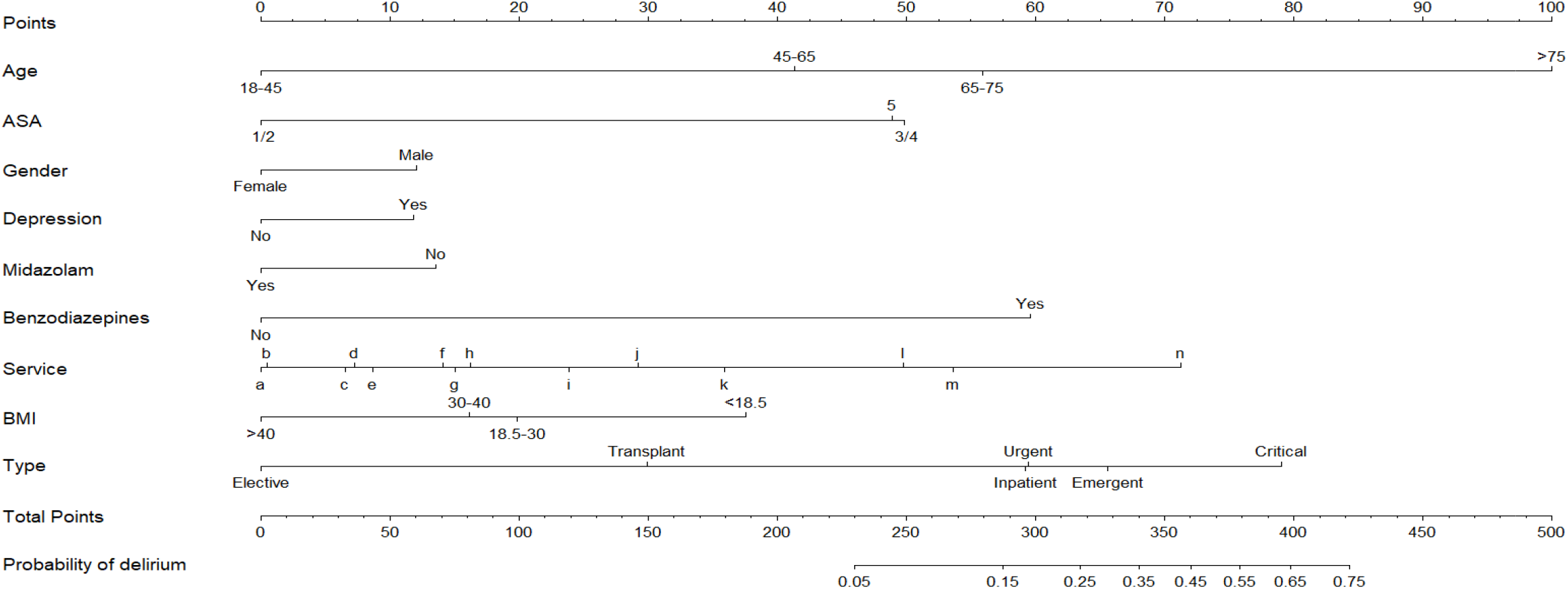
Nomogram Point to predict Individual risk for Postoperative Delirium (service legend a) Obstetrics and Gynecology, b) Gastroenterology, c) cardiac Surgery, d) Surgical Oncology, e) Otolaryngology, f) Other, g) Urology, h) General Surgery, i) Vascular Surgery, j) Thoracic Surgery, k) Plastic Surgery, l) Orthopaedics, m) Liver Transplant, n) Neurosurgery)

## Discussion

This is (to our knowledge) the largest retrospective cohort analysis of patients studied for POD to date. We examined nearly 33,000 patients aged >18 years old with an overall incidence of POD of 3.7%. The final multivariate model, containing nine features achieved an AUC of 0.85 (0.8-0.86) and was thus highly discriminating for POD. While many of our findings are similar to those of previous work in smaller studies on more focused populations, we report several differences. Like others, we found that POD was highly associated with advancing age, increased patient co-morbidities (ASA physical status), male gender, depression, high BMI, emergent cases, and invasive surgical procedures. However, we found no differences regarding type of anesthesia or if patients where premedicated with midazolam. The surgical services most likely where patients were more likely to develop POD where orthopaedics, liver transplant and neurosurgery.

Preoperative identification of high-risk patients for postoperative delirium is essential to allocate time and resources to implement targeted preventive strategies. As noted, this manuscript is not the first to perform a retrospective analysis of risk factors for POD.^27–29^ Medical literature contains many risk scores designs, each attempting to distill the likelihood of one of a variety of different outcomes. ^13,14–18^ However, to be easily calculated, were based on a limited number of highly predictive features. These scores evaluate disease status in broad strokes^30–33^ and often lack precision at the patient level – making them better suited to risk adjustment for populations than individualized risk prediction.^34^ Most of these studies were on smaller populations or focused on specific types of surgeries, or patient groups. For example, one important risk score tool for POD based on a large population (approximately 10,000 patients), recently developed by Whitlock et al^14^ accounts for precipitating factors solely by classifying surgical risk as moderate or high, and their risk score is unweighted and additive (i.e. summing the points for each risk factor), while clinical risk comes from the complex interplay of multiple factors. Our study differs in several ways. Firstly, rather than providing broad surgical groupings (low, medium and high risk) we stratified by surgical services while incorporating urgency as its own feature. Secondly, by directly harnessing EHR data we were able to incorporate a wider range of clinical co-morbidities hypothesized to be associated with POD including depression and BMI. Of note, the ability to increase the amount of data included in our model resulted in an AUC substantially higher than those reported previously. This gives further credence for the use of advanced modeling techniques, such as machine learning, that has been used to predict other conditions.^35–38^

In developing our model, we attempted to extract a wide variety of relevant data from the EHR to include as potential co-variates. Nonetheless, not all information was necessarily readily available. Important predictors, such as detailed information on cognition and sensory testing was not available. Further, previous studies (including several by our group) have repeatedly shown that EHR data is sparely populated and thus potentially inaccurate.^23^ We attempted to account for this by leveraging various types of EHR data to make more complete clinical phenotypes, however for patients less known to our system we may not have been sufficiently sensitive.

As important as the features that were associated with POD, are findings of features that were not associated with POD. In particular the use of midazolam as a preoperative anxiolytic has been the topic of much discussion for several years. In our cohort, this was actually protective of POD. However, it is critical to call out the limits of associations in a retrospective analysis such as this one. Given the awareness of theoretically associations between preoperative benzodiazepine use and POD, it is possible that there was significant selection bias in which patients were given midazolam preoperatively thereby affecting the results. Thus, these results must be interpreted carefully given the inability to address all possible confounders.

Importantly, we only included those patients who were screened for POD at least once during their hospitalization. Although, all patients admitted to the hospital should undergo delirium screening, it does not always happen. It is highly probable that patients at higher risk of delirium were screened more frequently introducing a selection bias to our cohort. In addition, previous work has shown that hyperactive delirium is more likely to be recognized by providers than hypoactive delirium. Our healthcare system has trained staff to recognize signs and symptoms of delirium. At least, for those patients that developed hyperactive delirium, it is highly likely that this information was introduced in the chart. Nonetheless, the exact rates reported in this manuscript may not be completely accurate and it is possible that the exact risk calculated based on our nomogram is not completely generalizable to patients who were not screened for POD. However, these broader trends and associations are likely correct and thus informative in creating a risk score.

POD is a multifactorial problem and until real-time clinical implementation of machine learning models become accurate and feasible, POD prediction that relies on weighted risk factors with an easy-to-use tool has the tremendous potential to improve predictive capacity and deployment of targeted interventions.

## Data Availability

All data produced in the present work are contained in the manuscript

## Authors’ contributions

SV: This author helped in study conception and design, data acquisition, data validation and interpretation and with manuscript drafting and revision. The author has approved the final version of the manuscript.

DC: This author helped in study design, data acquisition and data validation. The author has approved the final version of the manuscript.

TG: This author helped as methodology advisor, with statistical analyses, and with manuscript drafting and revision. The author has approved the final version of the manuscript.

IH: This author helped in study conception and design, data acquisition, data validation and interpretation, and with manuscript drafting and revision. The author has approved the final version of the manuscript.

## Acknowledgements

Emily Walters and Eilon Gabel for their assistance in data acquisition.

